# Parent attitudes about childhood vaccines: a vaccine hesitancy point prevalence study

**DOI:** 10.1101/2021.07.26.21261018

**Authors:** Sarah Marshall, Anne C Moore, Laura J Sahm, Aoife Fleming

## Abstract

**Objectives:** The aim of this study was to measure the prevalence of vaccine hesitancy using Parent Attitudes about Childhood Vaccines (PACV) survey regarding paediatric vaccines for their children, in a population of parents attending a STEM (Science, Technology, Engineering and Mathematics) outreach event in Cork, Ireland.

**Study design:** Cross-sectional survey study.

**Methods:** This study was conducted in November 2018 at the “Community Festival of Science” STEM event. Eligible attendees were invited to read the participant information leaflet, provide written informed consent, and complete the validated Parent Attitudes about Childhood Vaccines (PACV) survey. Each of the 15 PACV survey items was scored. A score ≥50 identified vaccine hesitant parents.

**Results:** A total of 105 parents participated in the study. A small number of participants (6.7%, n = 7) were identified as vaccine hesitant. There were no statistically significant differences between the vaccine hesitant and non-hesitant groups based on age, education, or number of children. Parents had concerns around vaccine side effects (36.2%, n=38) and vaccine safety (20%, n=21). Parents trusted the information they received on vaccines (85.6%, n=90) and 81.9% (n=86) believed that the vaccine schedule was good for their child.

**Conclusions:** The findings of this research indicate the presence of vaccine hesitancy in parents in Ireland regarding paediatric vaccines. Parents had concerns regarding vaccine side effects and the number of vaccines on the paediatric immunisation schedule. Further research is necessary to understand how these issues may contribute to vaccine hesitancy, and to develop evidence-based interventions to build on parents existing trust in the vaccination schedule.

**Key points:**

- A low level of vaccine hesitancy regarding childhood vaccinations was reported in a sample of parents at a science outreach event in Ireland using the PACV survey.
- Areas of parents’ concerns were around vaccine side effects and number of vaccines.
- Most parents trusted their doctors regarding vaccines, trusted vaccines information and followed the vaccine schedule.
- The PACV survey could be used as a screening tool to identify vaccine hesitant parents by healthcare providers and provide them with additional information and support, to support public health vaccination campaigns.

## Introduction

Vaccination has made the greatest contribution to global health of any human intervention, apart from the introduction of clean water and sanitation (1). Prophylactic vaccination of infants and children with several vaccines is the cornerstone of effective immunisation programmes against a variety of childhood diseases. Overall, it has been estimated that vaccines prevent almost six million deaths worldwide per annum (2). Despite these benefits, immunisation rates in many countries including Ireland, and for some vaccines, remain suboptimal in spite of vaccine availability (3). This has led to a series of potentially preventable disease (e.g. measles, pertussis) outbreaks internationally (4, 5). Waning vaccine confidence has taken a toll on immunisation programmes globally (6). While this reduction in confidence is multifactorial, a key contributory factor is vaccine hesitancy, recognised by the World Health Organisation as one of the greatest threats to global health in 2019 (7). Vaccine hesitancy refers to delay in acceptance or refusal of vaccines despite availability of vaccine services, is complex and context specific varying across time, place, and vaccines, and is influenced by factors such as complacency, convenience and confidence (8). This definition depolarises the pro- or anti-vaccine stance. In reality, vaccine-hesitant individuals are a heterogeneous group in a continuum, ranging from total acceptance to complete refusal (6). It is known that vaccine hesitancy is highly variable, and context specific, varying across time, place and vaccine involved (8, 9). It is a multi-layered phenomenon, related to prior beliefs about vaccines (10), perceived benefits of vaccines, attitudes towards vaccines (11), previous experiences with vaccines (12), socioeconomic status (13), number of children (14), and marital status (13). Given this heterogeneity, identifying vaccine-hesitant individuals in need of guidance is challenging. The Parent Attitudes about Childhood Vaccines (PACV) survey was developed to begin to address this challenge (15). It is a self-administered instrument and contains 15 items under three domains: behaviour, vaccine safety and efficacy, and general vaccine attitudes (15). The PACV survey has been validated to identify vaccine hesitant parents and to predict future vaccine refusal (16, 17). More recently, the survey has since been translated and tested in multiple languages (18-20), a short scale has been developed (21), and the instrument has been used in a wide variety of contexts (22). However, due to the context specificity of vaccine hesitancy, the reliability of the PACV survey should be assessed in different geographic areas and demographic samples of parents (17). Therefore, the aim of this study was to use the PACV to measure the prevalence of vaccine hesitancy in a population of parents attending the “Community Festival of Science”, a community outreach event in Cork, Ireland.

## Methods

This cross-sectional study is reported in line with the Strengthening Reporting of Observational Studies in Epidemiology (STROBE) statement. The study was conducted on the 18^th^ November 2018 at the “Community Festival of Science”: the finale of Cork Science Festival, involving highly interactive exhibitions and demonstrations. Cork Science Festival is a main partner of Science Week Ireland, one of the largest Irish STEM (Science, Technology, Engineering and Mathematics) engagement events. The study was approved by the Social Research and Ethics Committee, University College Cork (Log 2018-179). Any parent (over the age of 20 years) attending the event was eligible for participation in the study. Attendees were approached on an *ad hoc* basis throughout the event and invited to read the participant information leaflet about the study, to provide written informed consent, and to complete the PACV survey. The PACV is a validated survey and consists of 15 survey items under four content domains: immunisation behaviour (6 items), beliefs about vaccine safety and efficacy (8 items) attitudes about vaccine mandates and exemptions (1 item) and trust (3 items) (15). The survey was adapted for use in the Irish context e.g. highest education level reached (first, second, third or fourth level options in line with Irish context), and the replacement of the term “shot” in the survey questions with “vaccine”. In addition, questions pertaining to marital status, ethnicity and household income were removed. Some of the PACV pertain to the childhood vaccination schedule; the Irish schedule is provided as Supplementary information.

Each of the 15 PACV survey items was scored, in accordance with the original PACV system: hesitant responses are assigned a 2, ‘don’t know or not sure’ a 1 and non-hesitant responses a 0. Item scores were summed in an unweighted fashion to obtain a total raw score. The total raw score was then converted to a scale ranging from 0 (least hesitant) to 100 (most hesitant), using simple linear transformation, with a score ≥50 that identified vaccine hesitant parents, while a score <50 that identified non-hesitant parents (15). Continuous variables were described by medians and IQRs. Categorical variables were described by counts and percentages. Associations between categorical variables were investigated using Fisher’s Exact test. P values of <0.05 were considered statistically significant.

## Results

A total of 105 parents participated in the study. The survey was self-administered in less than five minutes and no issues were reported by participants with its completion. The most commonly reported age range was 40-49 years (n = 57, 54.3%), highest education level reached was third level (higher education in universities, institutes of technology and other colleges of education) (n = 79, 75.2%), and number of children was two (n = 53, 50.5%). A summary infographic of individual survey items and results depicting vaccine hesitancy is provided in Figure 1. The highest number of hesitant responses was associated with survey item *“How concerned are you that your child might have a serious side effect from a vaccine?”* with 36.2% of participants (n = 38) indicating they were somewhat concerned or very concerned (Figure 1). Conversely, the lowest number of hesitant responses was associated with survey item *“I am able to openly discuss my concerns about vaccines with my child’s doctor”*, with only 1.9% of participants (n = 2) reporting concern (Figure 1). In response to the item *“If you had another infant today, would you want him/her to get all the recommended vaccines?”*, 4.7% (n = 5) participants answered *“No”*, while 2.9% (n = 3) answered *“Don’t know”:* seven of these eight participants were identified as vaccine hesitant when scoring was complete. Most parents reported a non-hesitant belief that many of the diseases that vaccines prevent are severe (95.2%, n=100) and were accepting of vaccines in that they trust in vaccines information (85.6%, n=90).

**Figure 1.**
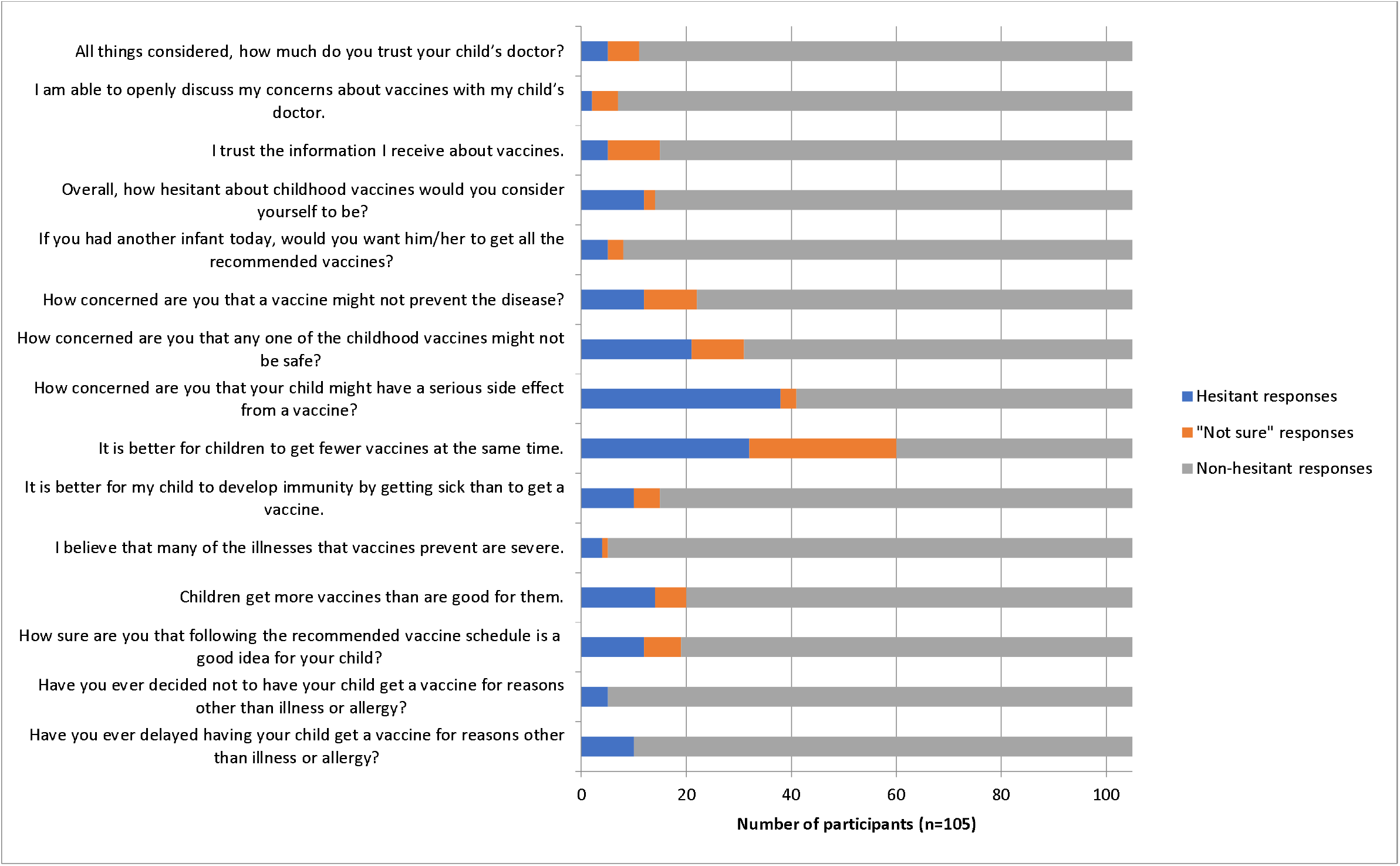
Responses to individual PACV survey items. A total of 105 parents participated in the study. The response by each parent to each of the 15 statements was classified as hesitant, “not sure” or non-hesitant.

Overall, 6.7% (n = 7) of participants were identified as vaccine hesitant, with a converted score ≥50 (Figure 2). The median (IQR) converted score was 10 (0, 20).

**Figure 2.**
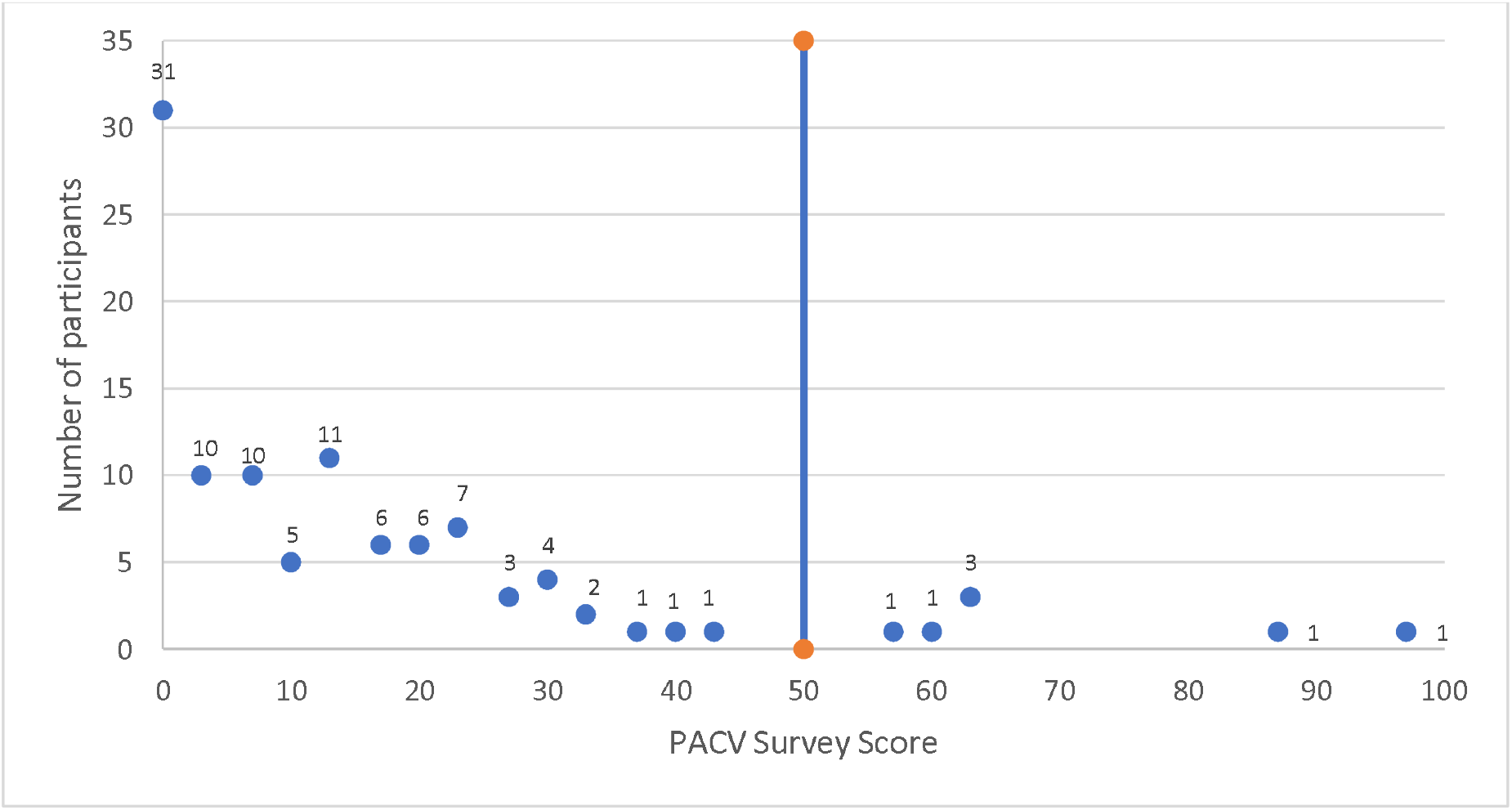
Participant PACV survey scores, with reference line at score 50.

There were no statistically significant differences between the vaccine hesitant and non-hesitant groups based on age (p=1.000), education (p=0.182) or number of children (p=1.000). Demographic information of those individuals identified as vaccine hesitant is presented in Table 1.

**Table 1.** Demographic information of vaccine hesitant participants.

| Participant | Age range | Highest | Number of |
| --- | --- | --- | --- |

|  | (years) | education level reached | children |
| --- | --- | --- | --- |
| 1 | 30-39 | Third | 2 |
| 2 | 40-49 | Second | ≥4 |
| 3 | 40-49 | Third | 1 |
| 4 | 40-49 | Third | 1 |
| 5 | 40-49 | Third | 2 |
| 6 | 40-49 | Third | 2 |
| 7 | 60-69 | First | 3 |

## Discussion

The aim of this study was to measure the prevalence of paediatrics vaccine hesitancy using the PACV survey, in a population of parents attending a STEM outreach event. The prevalence of vaccine hesitancy among participants was 6.7% and parents were mostly concerned about vaccine side effects and the number of vaccines on the childhood schedule. The PACV instrument has been administered in many regions worldwide, including Malaysia (18), the United States (23, 24), the UAE (25) and Italy (26), and the reported prevalence of vaccine hesitancy has been highly variable, ranging from 5.9% in a population of mothers in Washington State (24) to 26% of parents attending a paediatric emergency department of a tertiary hospital in Seattle (23). Here, we observed a low level of vaccine hesitancy prevalence in a cohort of parents attending a science festival.

This PACV instrument has been validated to predict future paediatric vaccine refusal. In this study, 87.5% of participants identified as vaccine hesitant according to the PACV survey (n = 6 of 7 in total) indicated that they would not, or were unsure whether they would consent to vaccination for future infants in their care. One further participant indicated that they were unsure, but were not identified as vaccine hesitant (PACV survey score = 30). This point prevalence study did not permit long term follow-up of future immunisation practices and it is known that intention alone does not necessarily predict future vaccine uptake: a disparity known as the intention-behaviour gap (27). However, this study suggests that the PACV survey may be used as a rapid screening tool to identify potentially vaccine hesitant parents as candidates for more intensive targeted vaccine education and decision support.

It is clear that healthcare providers such as doctors and nurses are perfectly positioned to screen patients and provide support. According to recent data collated by Wellcome Trust’s Global Health Monitor, which included the opinions of more than 140,000 people in over 140 countries, 93% of Irish participants reported that they trusted these healthcare providers and 85% trusted them most for medical and/or health advice (28). Similarly in this study, 94.3% of all participants (n = 99) agreed with the statement *“I am able to openly discuss my concerns about vaccines with my child’s doctor”*, and on a Likert scale ranging from 0 (do not trust at all) to 10 (completely trust), 89.5% of participants (n = 94) reported a score of ≥8 in response to the item *“All things considered, how much do you trust your child’s doctor?”* (**Figure 1**).

Factors such as number of children (14), and level of education (6) have been identified as determinants of vaccination. However, no such associations were identified in this population (). Fear of vaccine side effects has been consistently identified as a driver of vaccine hesitancy (29). This fear is often associated with new vaccines (30), novel delivery systems (31) or the sensationalist dissemination of vaccine misinformation. The purported links between MMR (measles, mumps, rubella) vaccination and autism (32), and HPV (human papillomavirus) vaccination and autoimmune conditions (33), have led to dangerous reductions in vaccine uptake. In this study, 36.2% of all participants reported concerns of side effects (**Figure 1**), but only 18.5% of these were identified as vaccine hesitant (according to the PACV survey), and the remaining participants (81.6%) indicated that they would consent to vaccination for future infants.

Vaccine hesitancy may also be driven by the belief that too many vaccines can “overwhelm the immune system” (34). In Ireland, the primary childhood immunisation schedule and school programme involve the administration of vaccines on 7 occasions from birth to approximately 13 years, and include both single and combination vaccines to protect against 13 diseases (**Supplementary Table 1**). Timing of the recommended schedule is important, taking into account waning of maternal antibodies and maturation of the immune system, susceptibility to the disease, and effectiveness and dosing of the vaccine (34) and research has demonstrated that a child’s immune system has enormous capacity to respond safely to multiple vaccines (35). However, in this study, 30.5% of participants felt that it is better for children to get fewer vaccines at the same time (**Error! Reference source not found**.). Similar to side effect concern, 78.1% of these participants declared they would vaccinate future children, and none were identified as vaccine hesitant. Therefore, it appears that parents will consent to vaccination in spite of these concerns but allaying these concerns is important in preventing their escalation, especially in the face of increased access to information sources of reliability (36). The essential role of healthcare providers in guiding vaccine decisions has been identified both in this study and in research conducted elsewhere, and effective interactions with providers can alleviate concerns of vaccine supportive parents and can motivate a vaccine hesitant parent towards acceptance (37). It has been shown that parents prefer strong, unambiguous recommendations from their provider (38). Development of continuous professional development (CPD) programmes to provide healthcare providers with information to adequately address vaccine concerns, and communication strategies to support providers in recommending vaccines with confidence would support HCWs efforts in this area. The importance of communication in addressing vaccine hesitancy has been identified (39), and future initiatives to guide healthcare providers interactions with parents on vaccines could address these concerns by means of a nationally implemented programme(40).

### Study strengths and limitations

A strength of the study is the use of a previously validated survey instrument (the PACV) to determine vaccine hesitancy in parents. A potential limitation of the study is the recruitment of participants from a science outreach event which is educational in nature, which may have increased the potential for increased vaccine acceptance. However, the vaccine hesitant parents had varying levels of education which may help to mitigate this risk. It has been shown in previous studies that education level does not predict vaccine hesitancy (6). In addition, there is the potential that self-selection bias impacted upon the results: those who are vaccine hesitant may have been less likely to participate in the survey upon invitation at the event. Conversely, the collection of data in an anonymous manner minimises the potential for social desirability bias. The study has a small sample size which limits the generalisability of the findings to the wider Irish population.

## Conclusions

The PACV survey was efficiently self-administered in a population of parents attending a STEM event. The prevalence of vaccine hesitancy was 6.7%. The survey results found that most parents reported as being non-hesitant on many areas such as trust in vaccines information and in the vaccination schedule. Some concern was expressed in the area of vaccine safety and side effect. The PACV survey may be used by healthcare providers to rapidly identify those parents, and their beliefs or concerns, in need of additional vaccine decision support and communication.

## Data Availability

Data from the study available as per manuscript.

## Conflict of interest

None declared.

## Funding

No funding was received for this study.

## Supplementary information

**Supplementary table 1:** Routine paediatric vaccination schedule in Ireland (Nov 2018). Reference: https://www.hse.ie/eng/health/immunisation/pubinfo/pcischedule/immschedule/

| Vaccine | Schedule |
| --- | --- |
| Primary immunisation schedule: |  |
| 6 in 1, PCV, MenB, Rotavirus | 2 months |
| 6 in 1, MenB, Rotavirus | 4 months |
| 6 in 1, PCV, MenC | 6 months |
| Measles Mumps Rubella (MMR), MenB | 12 months |
| Hib/MenC, PCV | 13 months |
| School Programme: |  |
| 4 in 1, MMR | Junior infants primary school (age 5-6 years) |
| Human Papillomavirus (for girls in 2018), Tdap (tetanus, diphtheria, pertussis), Meningococcal ACWY | First year secondary school (age 12-13) |

6 in 1: Diphtheria, Tetanus, (Pertussis), Hib (Haemophilus influenzae type b), Polio (inactivated), Hepatitis B.
PCV: Pneumococcal conjugate vaccine
MenB: Meningococcal B vaccine
MenC: Meningococcal C vaccine
Hib/MenC: Haemophilus influenzae type b, Meningococcal type b combined.
4 in 1: Diphtheria, polio, tetanus, pertussis.

## Notes

### Competing Interest Statement

The authors have declared no competing interest.

### Author Declarations

University College Cork Social Research Ethics Committee Log 2018-179

